# Phase 2b clinical study evaluating efficacy of RTS,S/AS01_E_ in *Plasmodium falciparum*-exposed Kenyan adults treated with antimalarial chemopreventive drugs

**DOI:** 10.1101/2025.01.10.25320353

**Authors:** Nathanial K. Copeland, Lucas Otieno, June Doryne Otieno, Solomon Otieno, Salome Chira, Karen Ivinson, Irene Onyango, Ruth Wasuna, Hoseah Akala, Amos Onditi, Peter Sifuna, Ben Andagalu, Roselyne Oyugi, Mary Omondi, Stellah Amoit, Emily Locke, Scott Gregory, Elke S. Bergmann-Leitner, Hema Pindolia, Mike Raine, Chris Gast, Laina D. Mercer, John J. Aponte, Marc Lievens, Christian F. Ockenhouse, Cynthia K. Lee

**Affiliations:** Kombewa Clinical Research Centre, Kenya Medical Research Institute/US Army Medical Research Directorate-Africa, Kisumu, Kenya; Center for Vaccine Innovation and Access, PATH, Washington DC, USA; Seattle, USA; Nairobi, Kenya; and Geneva, Switzerland; Walter Reed Army Institute of Research, Silver Spring, MD, USA; GlaxoSmithKline, Wavre, Belgium; Human Immunology Laboratory, International AIDS Vaccine Initiative, London, UK

**Keywords:** Malaria vaccine in African adults, vaccine-induced antibodies and infection, RTS, S/AS01

## Abstract

**Background:** RTS,S/AS01 vaccine efficacy (VE) was previously shown to be lower in African adults than in malaria-naïve US adults, potentially due to concurrent *Plasmodium falciparum* (*Pf*) infections. We investigated whether treatment of infection prior to vaccination would lead to improved VE and immunogenicity.

**Methods:** A Phase 2b study in Kenyan adults evaluated the efficacy of RTS,S/AS01_E_ in conjunction with antimalarial chemopreventive drugs. Participants, grouped by baseline presence or absence of *Pf* infections, were randomized to receive RTS,S/AS01_E_ or rabies vaccine. Four groups received antimalarial drugs prior to immunization and were followed for six months to assess *Pf* infection. We included an additional group of adults not treated with antimalarial drugs for immunological assessment.

**Results:** VE (RTS,S/AS01_E_ versus rabies vaccine) was 34.8% (8.9%, 53.4%) and −24.0% (−97%, 22.4%) in baseline *Pf*-positive and baseline *Pf*-negative participants, respectively. In RTS,S/AS01_E_ recipients, there were no statistical differences in anti-circumsporozoite (CS) antibody titers in baseline *Pf-*positive or *Pf*-negative participants, or in susceptibility to infection during the post-vaccination follow-up period. Drug treatment did not improve anti-CS antibody titers.

**Conclusions:** Treating *Pf* infections during vaccination does not result in increased VE. Anti-CS antibody responses to vaccination do not differ with baseline *Pf* infection status, drug treatment, or susceptibility to *Pf* infections.

**Clinical Trial Registration:** NCT04661579; PACTR202006896481432

## INTRODUCTION

The World Health Organization prequalified and recommended the RTS,S/AS01_E_ vaccine to protect against clinical malaria disease in young children living in sub-Saharan Africa(1,2) More than 6 million doses were administered by national immunization programs in Ghana, Kenya, and Malawi in a large-scale pilot program (3) In order to inform whether circumsporozoite (CS)-based *Plasmodium falciparum (Pf)* vaccines could have a role in parasite elimination, it is critical to evaluate whether they protect against infection in all age groups that serve as reservoirs of parasites contributing to ongoing transmission.

Proof-of-concept evidence for RTS,S was initially derived in malaria-naïve adults in the United States using a controlled human malaria infection (CHMI) challenge model where vaccine efficacy (VE) of 52-63% was achieved with RTS,S/AS01_B_ formulation in moderate-sized trials (4,5). However, when RTS,S was tested in Kenyan adults living in high malaria transmission regions (6) VE after 9 weeks of active and passive case detection against smear-positive *Pf* was only 29.5% (95% confidence interval [CI]: −15.4-56.9, *p* = 0.164).

It is recognized that in immunologic hyporesponsiveness, T cell exhaustion and B memory cell dysfunction, as measured by decreased immune response to pathogens, is influenced by concurrent asymptomatic sub-microscopic *Pf* parasitemia (7–13). This immunologic hyporesponsiveness may impede the development of protective immune responses following immunization. Higher VE achieved with adjustments in vaccination schedule and dosage (87% in fractional-dose regimen compared to 63% for the standard-dose regimen) in more recent CHMI studies (5,14,15) provided an added incentive to test the delayed fractional-dose regimen in African adults. This Phase 2b trial tested the efficacy and immunogenicity of the fractional-dose regimen of RTS,S in adults with or without baseline malaria infections and treated with antimalarial chemotherapy.

## METHODS

The protocol for this Phase 2b, randomized, open-label, controlled, single-center study was approved by the Kenya Medical Research Institute (KEMRI) Scientific and Ethics Review Unit and the Walter Reed Army Institute of Research Institutional Review Board. The trial was conducted in accordance with ICH guidelines and Good Clinical Practice. Written informed consent was obtained from each subject before study initiation.

### Study Design

We enrolled five groups of participants who were randomized by the presence or absence of PCR-positive *Pf* parasitemia at baseline to receive antimalarial treatment, RTS,S/AS01_E_, or a comparator rabies vaccine (Figures 1, 2, Supplementary Tables S1, S2, S3). The primary objective of the study was to assess VE in a cohort <u>positive</u> for *Pf* by PCR at baseline (Groups 1 and 4), the secondary objective was to assess VE in a cohort <u>negative</u> for *Pf* by PCR at baseline (Groups 2 and 5). Both cohorts were treated with antimalarials prior to each vaccination and VE was assessed by time to first *Pf* infection as measured by PCR. Participants in Group 3, included to serve as immunologic controls, were <u>positive</u> for *Pf* by PCR at baseline and received RTS,S/AS01_E_ but not antimalarial drugs.

**Figure 1.**
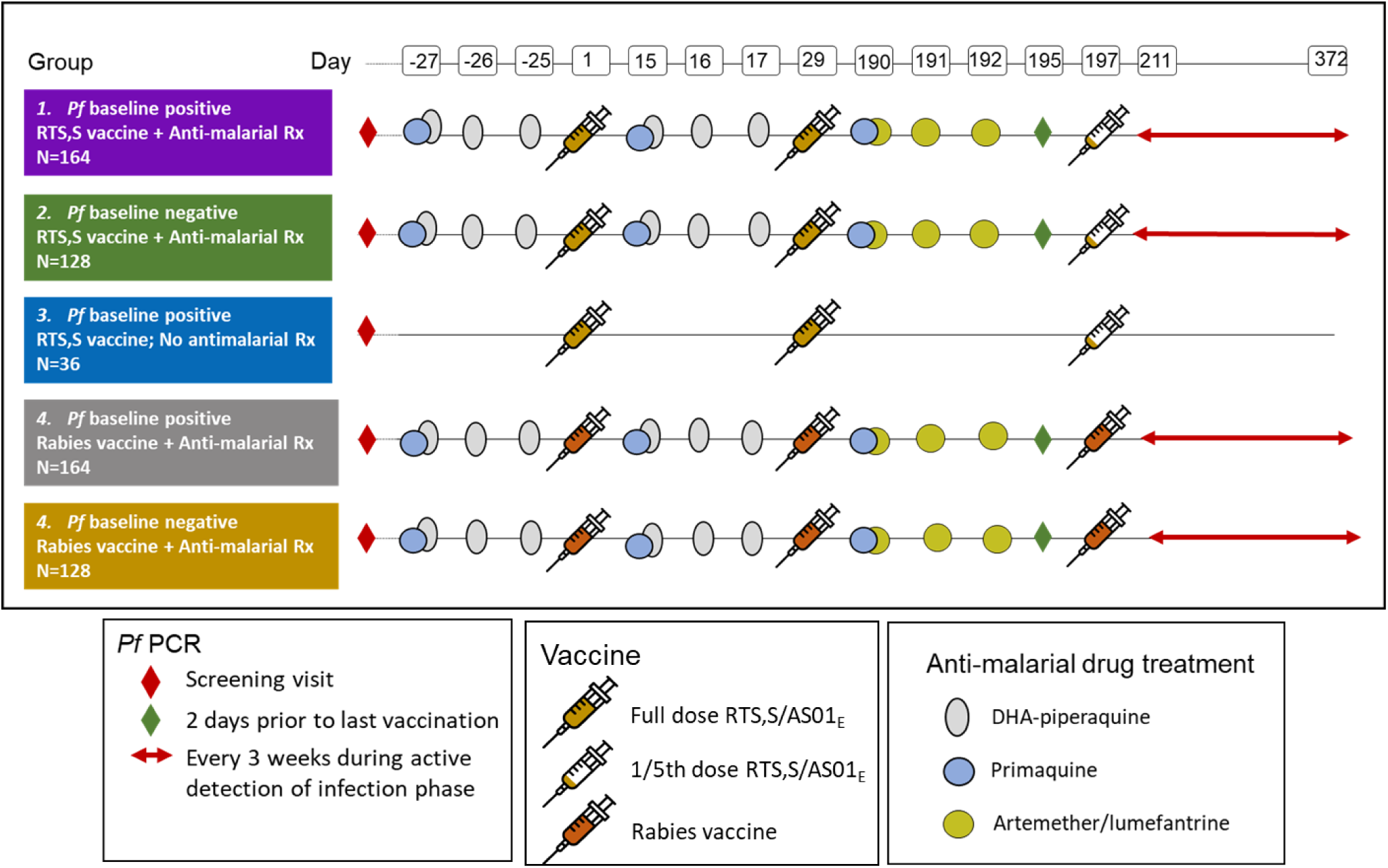
Study design. Abbreviations: ADI, active detection of infection; Fx, fractional; *Pf*, *Plasmodium falciparum*.

**Figure 2.**
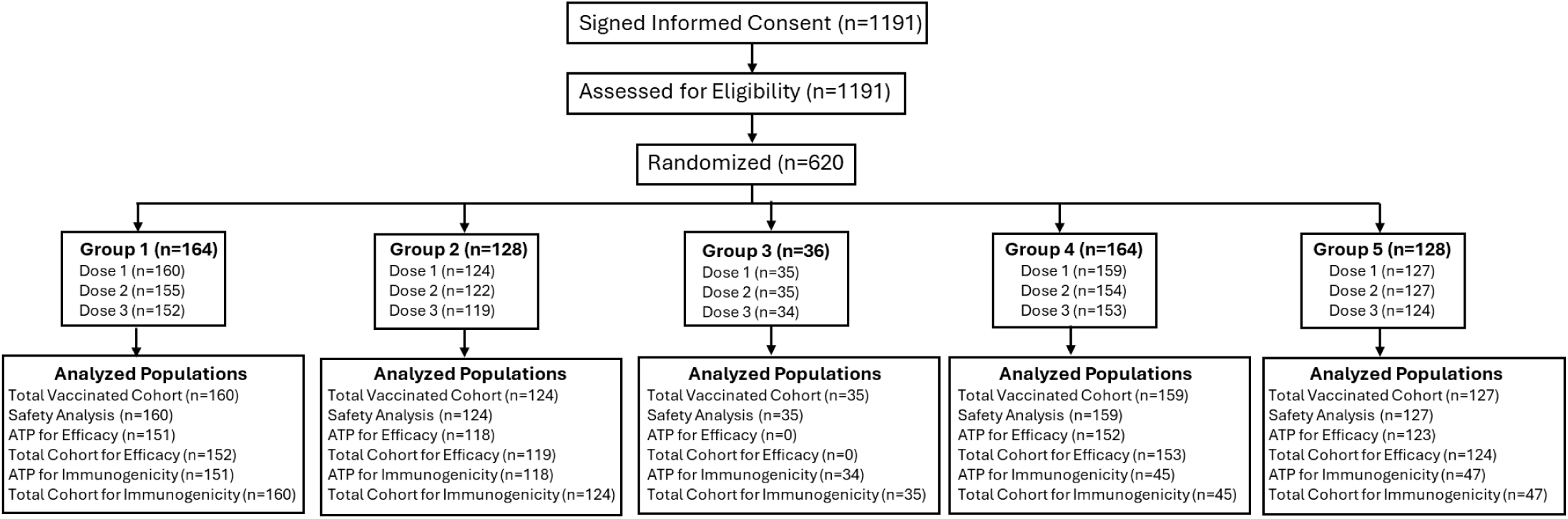
Consort diagram.

### Participants

The trial was conducted at the KEMRI-Walter Reed Project’s Kombewa Clinical Research Center in healthy adults recruited from the village of Seme and half of the Kisumu West Sub-Counties of Kisumu County, Kenya. Eligible participants included males and nonpregnant females aged 18–55 years with no serious acute or chronic illness as determined by medical history, physical examination, record review, or laboratory screening tests. Participants with below Stage 3 HIV, who were otherwise healthy and on antiretroviral therapy, were included. Participants were stratified by HIV and *Pf* infection statuses and randomized to one of five study groups (Figure 2, Supplementary Table S1).

### Study Treatments

Antimalaria drugs dihydroartemisinin (40 mg) and piperaquine tetraphosphate (DHA/Pip; 320 mg); artemether (20 mg) and lumefantrine (A/L; 120 mg); and primaquine phosphate tablets (26.3 mg) were administered to participants in Groups 1, 2, 4, and 5 (Figure 1, Supplementary Table S2). DHA/Pip was chosen for its extended half-life pharmacokinetic properties to provide “parasite-free” status for an extended period between immunizations. A/L was selected to be given prior to the third vaccine dose for its short half-life to clear any blood-stage parasites and establish a clean baseline for determination of VE during the active detection of infection (ADI) period of observation. Low-dose primaquine (15 mg) was used to clear circulating mature, sexual-stage gametocytes that may interfere with the PCR assay.

### Study Vaccines and Vaccination

RTS,S/AS01_E_ is manufactured by GlaxoSmithKline (GSK, Rixensart, Belgium). The pediatric formulation for vaccine doses 1 and 2 contained 25 μg of RTS,S and an adjuvant system AS01^E^ (25 μg of MPL, 25 μg of QS21, and liposomes) in 0.5mL. At dose 3, participants were administered a fractionated dose of RTS,S/AS01_E_ (0.1 mL; one-fifth of the antigen and adjuvant dose). Participants randomized to the control comparator group received three equal doses of rabies vaccine (Abhayrab®, Human Biologicals Institute, Andra Pradesh, India). All vaccines were administered intramuscularly in the deltoid muscle of the arm on a 0-, 1-, and 7-month schedule.

### Safety Assessment

Solicited local and systemic adverse events (AEs) in the reactogenicity cohort following the administration of each dose of vaccine were recorded for the first 50 participants in Groups 1 and 2 and for all participants in Group 3. Unsolicited AEs occurring within 28 days following administration of each dose of vaccine and, where applicable, courses of per protocol scheduled antimalarial treatment were recorded. Serious adverse events (SAEs) and pregnancies in participants were recorded until study end.

### Efficacy Assessment

One week prior to dose 3 all participants in Groups 1, 2, 4, and 5 were presumptively treated with A/L and primaquine prior to the start of ADI. Participants in Group 3 did not receive antimalarial treatment, were not included in the ADI phase of the study, and were followed for immunogenicity only. Efficacy evaluation included both ADI with blood draws every three weeks and passive case detection in all participants presenting with symptoms consistent with malaria. The primary analysis, defined as the time to first *Pf* infection, detected by a *Pf*/pan*-Plasmodium* 18S rRNA PCR (16), was conducted on the According-to-Protocol (ATP) cohort. Based on previous experience with assessing gametocyte carriage (17), a cycle threshold of 31, equivalent to <1.28 parasites/µL was used as the cut-off for positive result. A cross-sectional prevalence assessment of *Pf* parasitemia by PCR at study end (approximately 500 days after study start) was also conducted.

### Immunogenicity Assessments

Antibody levels and avidity against the central repeat region (NANP) and the carboxy-terminal domain of the CS protein region were measured using validated standard enzyme-linked immunosorbent assays (ELISAs) performed at the ImmunCore/Malaria Serology Laboratory at Walter Reed Army Institute of Research (14). Antibody concentrations against the hepatitis B surface antigen (HBs) were performed at the Human Immunology Laboratory at the International AIDS Vaccine Initiative using an in-house validated assay according to ICH guideline Q2(R1) (18), based on the Monolisa™ Anti-HBs PLUS kit (BIO-RAD, Hercules, California, USA).

Anti-rabies immunoglobulin G (IgG) titers were performed at Kansas State Veterinary Diagnostic Laboratory using a validated ELISA based on the Platelia Rabies Kit II (BIO-RAD, Hercules, California, USA).

### Statistical Analyses

The statistical analyses were conducted following the principles specified in ICH Topic E9 (19) In accordance with the predefined analysis plan, the analyses were performed after database lock using SAS 9.4 (SAS Institute Inc., Cary, North Carolina, USA).

The analysis of efficacy endpoints for the primary and secondary objectives and endpoints were defined by time to a positive PCR for the presence of *Pf* parasites meeting the definition of an event, from a blood sample collected at any time during the ADI phase of the trial in the ATP for efficacy cohort. The impact of the event-driven study design was to continue to follow up until 92 first infection events were observed in aggregate between Groups 1 and 4, and 72 first infection events were observed in the aggregate of Groups 2 and 5, up to a maximum of 12 months of ADI. The primary and secondary endpoints were assessed using Cox proportional hazards regression (time to first PCR-positive infection) with a covariate for group assignment (Groups 1 and 4 for primary endpoint, and Groups 2 and 5 for secondary endpoint).

The primary immunogenicity analysis was based on the ATP for immunogenicity cohort. A secondary analysis, based on the total vaccinated cohort, was performed to complement the ATP analysis.

For the safety analysis, data from all participants in the safety analysis population were included. All analyses were descriptive and performed on the intent-to-treat set that included all participants who received ≥1 dose of a study vaccine.

## RESULTS

### Demographic Characteristics

From November 2020 to May 2021, 1191 volunteers (>98% Luo ethnicity) were assessed for eligibility to participate, and 620 participants were randomized into the study groups in accordance with their baseline *Pf* status (Figure 2, Supplementary Tables S1, S2). Overall, 605 participants were administered study vaccine. Reasons for participants not receiving study vaccine included pregnancy, noncompliance with study procedures, withdrawal of consent, alcohol abuse, and personal reasons. Of the 605 participants who received study vaccine, 582 (93.9%) participants received all scheduled vaccinations, and 539 (89.1%) participants completed the study. The mean age (SD) was 32.91 (±8.19) years; 323 [53.4%]) were female and 282 [46.6%] were male; 103 (17%) participants were positive for HIV infection; 507 (83.8%) participants owned a bed net and slept under it daily; and 546 (93.5%) participants were fully compliant with antimalarial treatment (Supplementary Table S3).

### Vaccine Efficacy

The ADI phase commenced 14 days after the third dose of vaccine was administered on Day 197 in participants from Groups 1, 2, 4 and 5. All individuals in the four groups had negative *Pf* PCR test results on the day they were administered vaccine dose 3 and five days after completing A/L plus primaquine regimen.

Participants in Groups 2 and 5 (negative baseline parasitemia) were recruited more quickly than those in Groups 1 and 4 (positive baseline parasitemia), so the period at risk for new *Pf* infection during the ADI phases of the two cohorts largely overlapped but were not identical. In the control rabies vaccine groups, the attack rate during the ADI phase of the study was 1.48 events/person years at risk (PYAR) in Group 4 and 0.62 events/PYAR in Group 5 (Table 1).

**Table 1.**
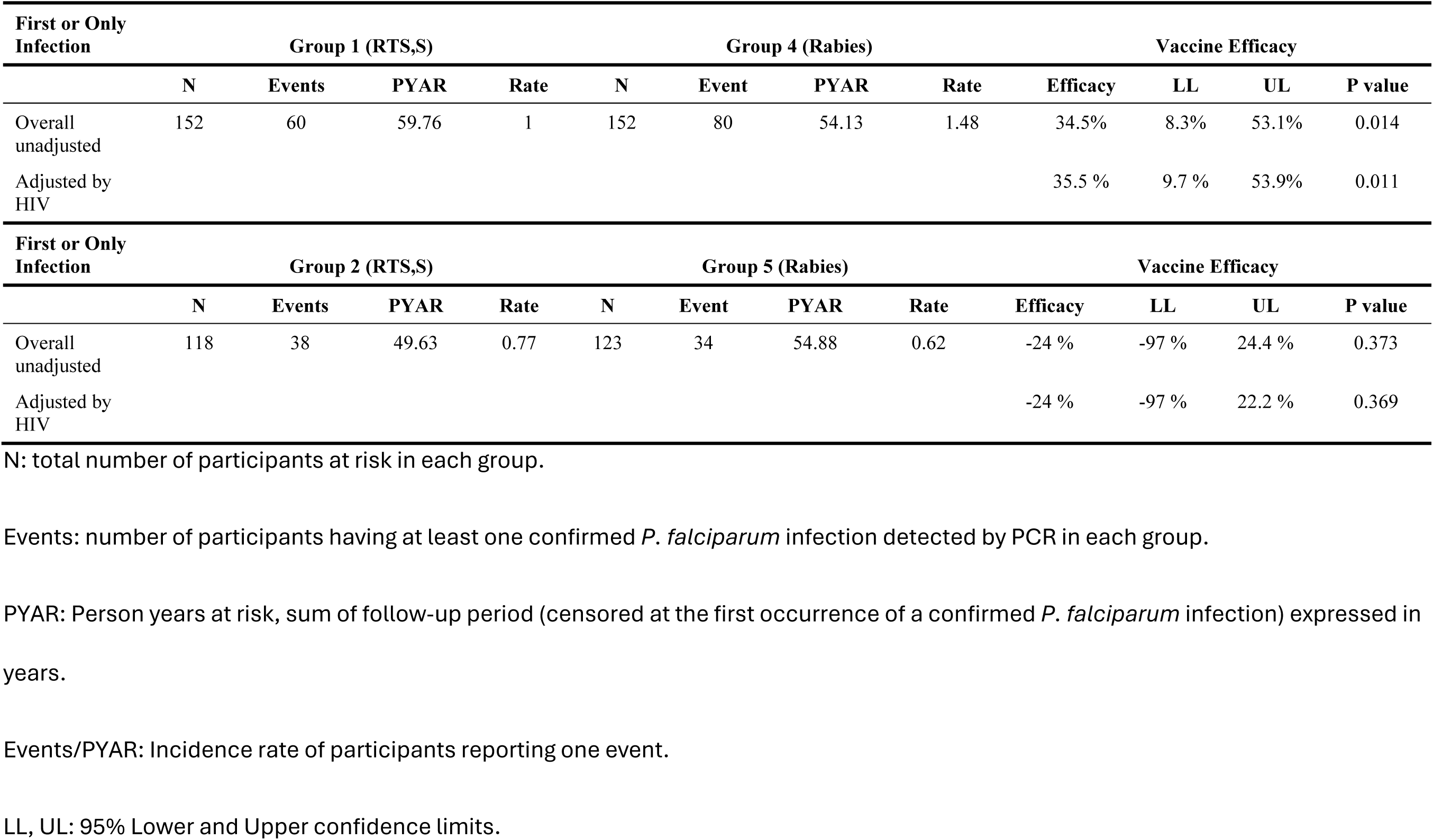
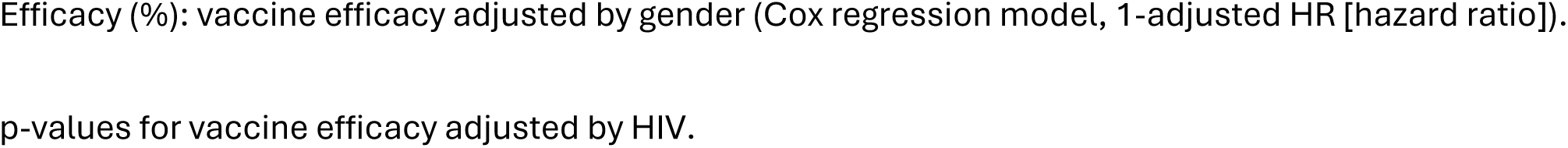
Vaccine efficacy, First infection events of *P. falciparum* during the active detection of infection period by HIV status and overall for Groups 1 and 4 and for Groups 2 and 5 (Total Cohort for Efficacy)

#### VE against incident pf infection (RTS,S/AS01_E_ versus rabies vaccine) in baseline Pf PCR positives

The primary objective was to determine VE in those who were *Pf* PCR positive at baseline. Overall, in the ATP cohort during the six-month period following the last dose of the vaccine, 60 PCR-detectable infection events were recorded for Group 1 and 80 events were recorded for Group 4 during ADI. The person-years at risk (PYAR) was 59.53 for Group 1 and 53.67 for Group 4, and the incidence rate was lower for Group 1 (1.01 events/PYAR) compared to Group 4 (1.49 events/PYAR). When compared to the rabies vaccine, the overall estimated VE of RTS,S/AS01_E_ was 34.8% (95% CI; 8.9-53.4%; *p* = 0.012) (Table 1). Similar results were observed for the total cohort for efficacy (results not shown). When adjusted by HIV status, the estimated VE was 35.9% (95% CI; 10.3-54.2%; *p* = 0.009). HIV status had no statistically significant impact on VE (p = 0.247). No other potential confounding covariates at baseline that reached the cut-off of *p* < 0.1 were included in the adjusted model. The time to PCR positive parasitemia is shown in a Kaplan-Meier survival plot (Figure 3A) with evidence for a difference in event-free survival time between Groups 1 and 4.

**Figure 3.**
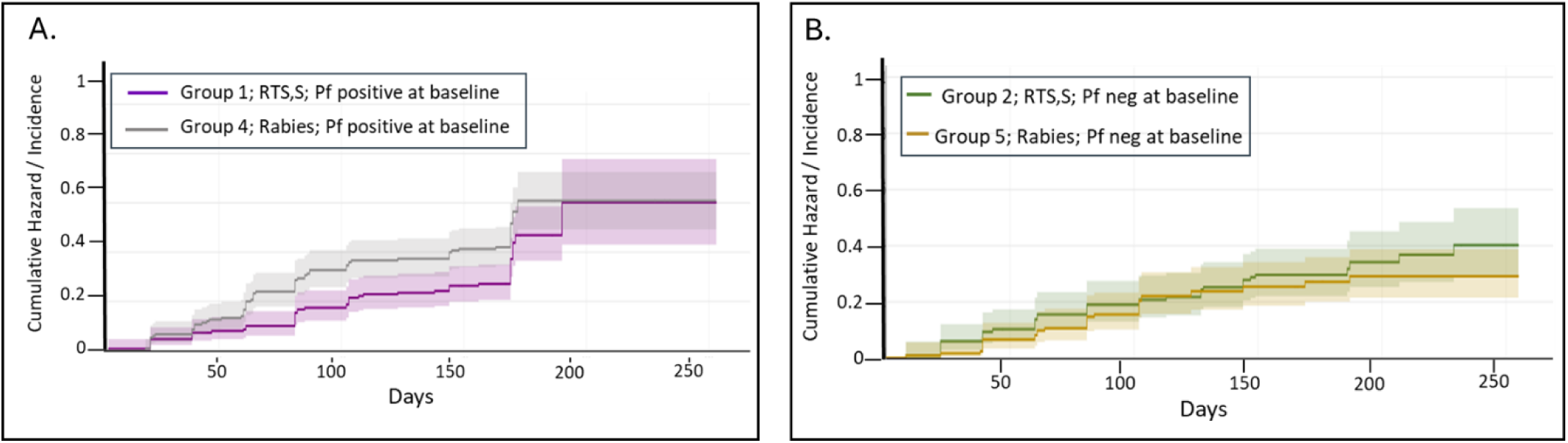
Vaccine efficacy shown as Kaplan-Meier survival plots comparing the event-free survival between A. Groups 1 and 4 (primary endpoint), and B. Groups 2 and 5 (secondary endpoint)

#### VE against incident pf infection (RTS,S/AS01_E_ versus rabies vaccine) in baseline Pf PCR negatives

The secondary objective was to determine VE in those who were *Pf* PCR negative at baseline. Overall, in the ATP cohort, 38 events were recorded for Group 2 and 34 events were recorded for Group 5. The PYAR was 49.63 for Group 2 and 54.88 for Group 5. The incidence rates for Group 2 and 5 were similar (0.77 and 0.62 events/PYAR, respectively). The unadjusted estimated VE was not significant, −24% (95% CI; −97-22.4; *p*=0.373) (Table 1) and was similar when adjusted by HIV status. Similar results were observed for the total cohort for efficacy (results not shown). When adjusted by HIV status, the estimated VE was −24% (95% CI; −97-22.2%; *p* = 0.369). The time to PCR positive parasitemia is shown in a Kaplan-Meier survival plot (Figure 3B) with no evidence of a difference in survival times between Groups 2 and 5.

### Immunogenicity

Anti-CS NANP repeat and C-terminus (C-term) geometric mean IgG titers and antibody avidity were measured in all participants immunized with RTS,S/AS01_E_ (Groups 1, 2, and 3). Prior to RTS,S/AS01_E_ vaccination, Groups 1, 2, and 3 had approximately 78%, 67%, and 74% of participants, respectively, with anti-CS NANP repeat antibody responses and 89%, 78%, and 94% of participants, respectively, with anti-CS C-term antibody responses above the lower limit of quantification.

RTS,S/AS01_E_ was immunogenic in all groups based on geometric mean titer (GMT) to both the NANP repeat and the C-term domain measured at baseline and after each immunization. There was no significant effect on anti-CS antibody titers to either the NANP repeat or the C-term domain post-immunization in individuals who were baseline positive (Groups 1 and 3) or negative (Group 2) for *Pf* infection (Figure 3). The anti-NANP antibody titers decreased over time but increased after the fractional dose at month 7, albeit not to the levels achieved after the second dose. Despite parasite clearance (Group 1) or chemoprevention (Group 2) with antimalarial medications, the antibody kinetics were comparable among all groups (Figure 4). There was no difference in anti-NANP (Supplementary Table S5) or anti-C-term (Supplementary Table S6) avidity indices across Groups 1, 2, and 3.

**Figure 4.**
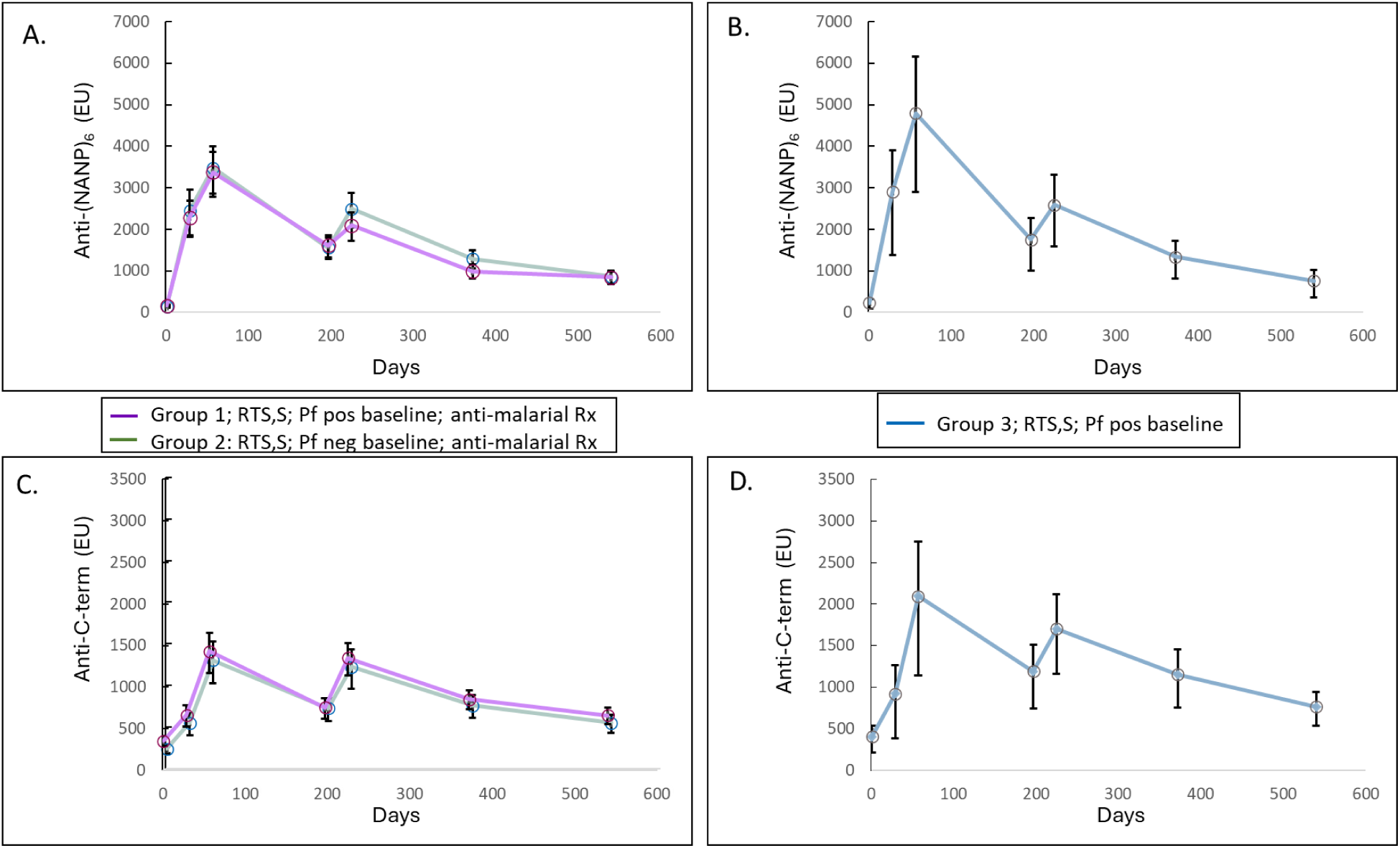
Anti-circumsporozoite NANP and C-term antibody kinetics. A. Anti-NANP repeat titers from Groups 1 and 2 who received RTS,S and antimalarial medications compared to, B. titers from Group 3 who received RTS,S but no antimalarial medications. C. Anti-C-term titers from Groups 1 and 2 who received RTS,S and antimalarial medications compared to, D. titers from Group 3 who received RTS,S but antimalarial medications. Abbreviations: EU, ELISA units, equivalent to Geometric Mean Titers; *Pf*, *Plasmodium falciparum*; Rx, medication.; *Pf*, *Plasmodium falciparum*; Rx, medication.

The individual post-dose 3 anti-NANP and anti-C-term antibody titers in participants immunized with RTS,S/AS01_E_, presented in boxplots and reverse cumulative distribution curves showed no differences between adults with or without *Pf* parasites in their blood at baseline (Figure 5). Nor were there differences in anti-CS antibody responses post-immunization in HIV-positive or HIV-negative participants in either Group 1 or 2 (Supplementary Figure S1).

**Figure 5.**
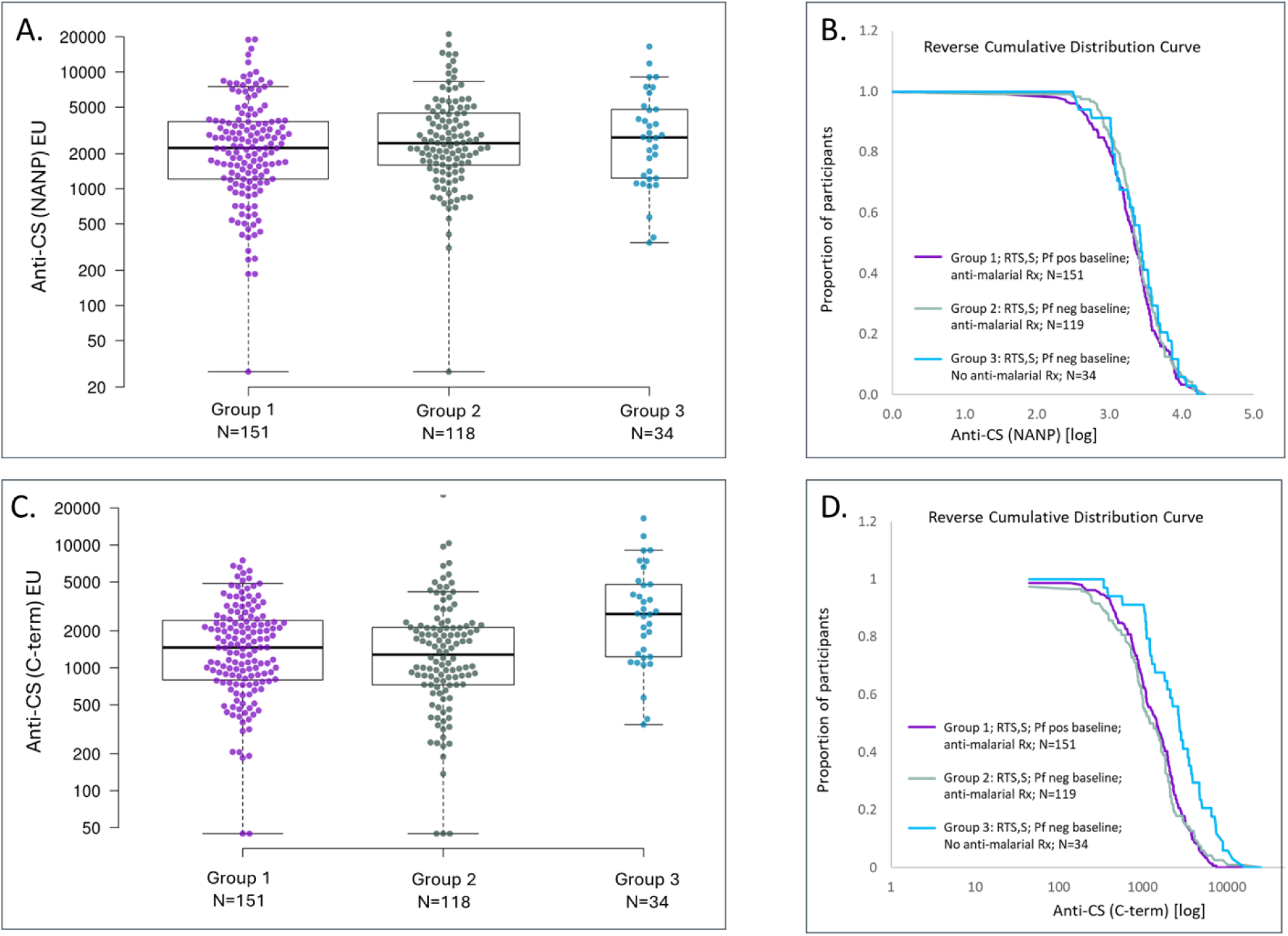
Anti-circumsporozoite NANP and C-term titers in Groups 1, 2,3 at post-dose 3 (day 225). Anti-NANP titers shown as scatter plots (A) and reverse cumulative distribution curve (B). Anti-C-term titers shown as scatter plots (C) reverse cumulative distribution curve (D).. Abbreviations: CS, circumsporozoite; EU, ELISA units, equivalent to Geometric Mean Titers; *Pf*; *Plasmodium falciparum*; Rx, medication.; *Plasmodium falciparum*; Rx, medication.

Ancillary analyses were performed to evaluate the association of anti-CS antibodies with VE. Anti-CS GMTs measured one month post-dose 3 in Groups 1 and 2 were analyzed with respect to whether they experienced a *Pf* PCR positive event during the six-month ADI period. Both anti-CS NANP and C-term antibody GMTs were equivalent in noninfected compared to infected participants (Figure 6).

**Figure 6.**
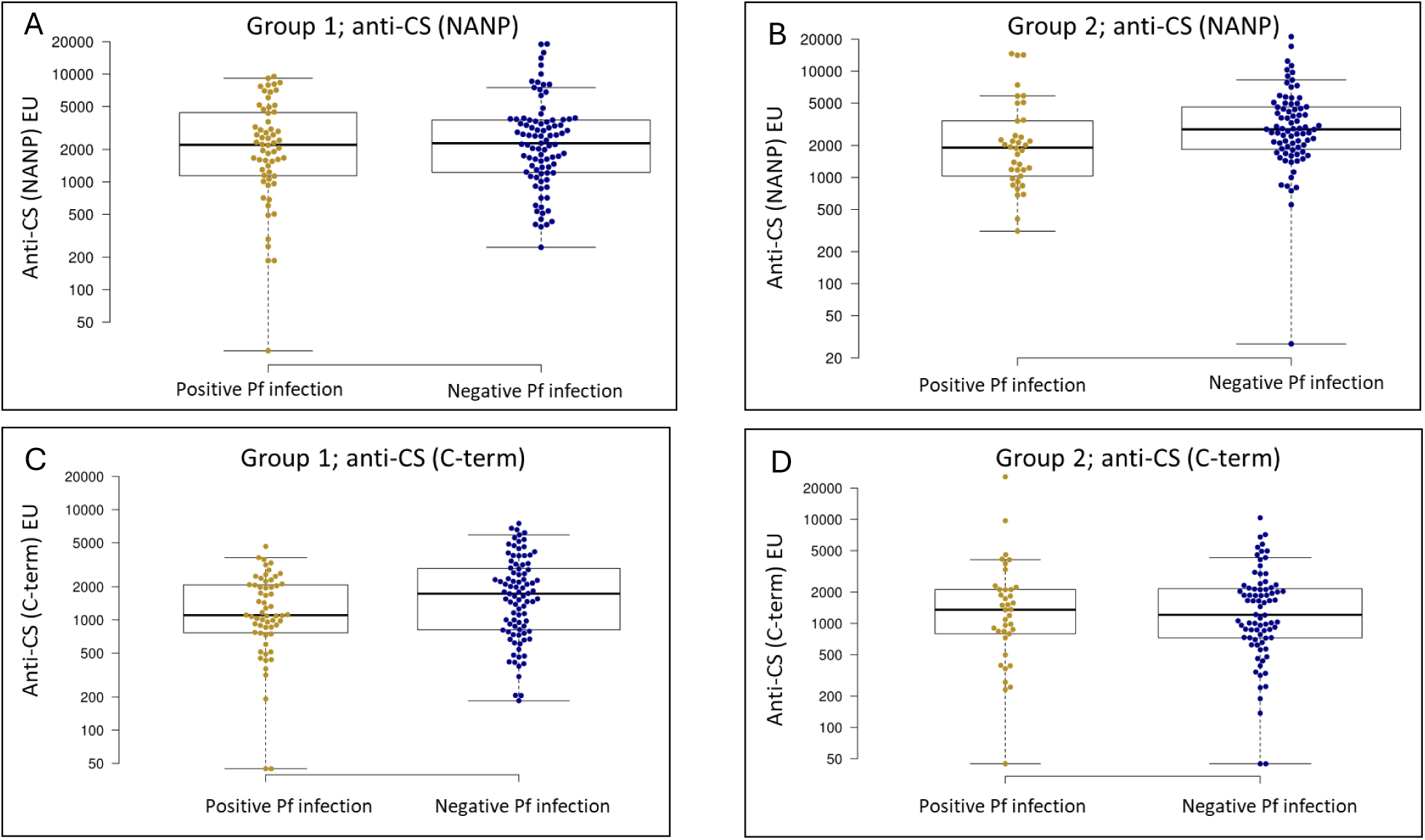
Anti-circumsporozoite NANP and C-term titers post-dose 3 (day 225) stratified by *Pf* infection outcome during the active detection of infection phase. A. Anti-NANP titers within Group 1 (A) and Group 2 (B). Anti-C-term titers within Group 1 (C) and Group 2 (D). Abbreviations: CS, circumsporozoite; EU, ELISA units, equivalent to Geometric Mean Titers; *Pf, Plasmodium falciparum*.

Antibody titers against hepatitis B surface antigen, a component of the RTS,S antigen, were similar in baseline parasitemic versus non-parasitemic individuals (Group 1 versus 2) and in individuals who received antimalarial drug treatment or not (Group 1 versus 3) (Supplementary Figure S2). Consistent with prior studies, a marked increase in antibody levels from baseline to protective thresholds of anti-HBs titer was achieved in all RTS,S-vaccinated groups.

There was no significant difference in anti-rabies IgG titers in participants who were parasitemic at baseline (Group 4) compared to participants who were not parasitemic at baseline (Group 5) (Supplementary Figure S2).

### Safety

The RTS,S/AS01_E_ regimen was well-tolerated. The number of participants per group and the number of local and systemic elicited AEs, the number of unsolicited AEs, and the number of SAEs is shown in Supplementary Table 4.

The proportion of participants experiencing local and systemic solicited AEs was similar across groups, and most participants had AEs of mild severity. The most frequently reported local solicited AE was pain at the site of injection. The most frequently reported systemic solicited AE was headache. Unsolicited AEs were mild in severity in most participants, and all AEs were unrelated to study treatment.

Overall, 13 SAEs were reported in 10 participants (3 participants in Group 1; 2 participants in Group 2; 1 participant in Group 3; 1 participant in Group 4; 3 participants in Group 5). One SAE resulted in death (alcoholic coma), and one led to discontinuation. No SAE was considered related to study treatment.

Seven participants became pregnant during the immunization phase of the study (5 received at least one dose of RTS,S/AS01_E_ and 2 received at least one dose of rabies vaccine) and were withdrawn from any further immunizations but were followed for safety until delivery or termination of the pregnancy. There were no pregnancies in women who completed all vaccinations. There was one participant in Group 3 whose pregnancy ended in spontaneous abortion, determined as not related to vaccination.

## DISCUSSION

The introduction of two malaria vaccines, RTS,S/AS01_E_ and R21/Matrix-M, to prevent clinical malaria disease in young children in sub-Saharan Africa has been a welcomed supplemental tool. Studies in African adults are necessary to assess how these and future vaccines may be harnessed to accelerate *Pf* elimination in populations with different age structures, different histories of exposure and acquired immunity, and different epidemiological contexts spanning low to high malaria transmission intensities.

This study used the pediatric formulation of RTS,S/AS01_E_, containing half the active ingredients of the adult formulation RTS,S/AS01_B_, as a prototype for CS-based vaccines, which was administered using a delayed fractional third-dose regimen. We chose this regimen as CHMI studies demonstrated superior efficacy with a delayed fractional third-dose regimen (5) and comparable efficacy using the pediatric formulation in adults (14). Moreover, data from field trials in young African children showed no reduced VE in children receiving a delayed fractional-dose regimen over the standard full-dose regimen during 12 months following the third dose (20).

This study was designed to support eventual use-case scenarios which combine a vaccine that prevents infection with antimalarial medications that clear or prevent parasitemia. Recent studies combining RTS,S/AS01_E_ with seasonal malaria chemoprevention in young African children resulted in a synergistic effect in averting clinical malaria episodes (21). Additionally, there is evidence that acute clinical malaria and/or asymptomatic malarial parasitemia may suppress the induction of protective antibody-mediated immune responses to vaccines (7–13). We hypothesized that antimalarial chemotherapy could reverse this immunosuppression and enhance the overall effectiveness of a vaccine. We also expected that VE would be higher in RTS,S/AS01_E_-immunized individuals who were *Pf*-negative at study start. Our study results showed that neither of these hypotheses were correct. Antimalarial drug treatment did not substantively improve VE to the levels achieved in CHMI studies in *Pf*-naïve adults, nor in comparison to the earlier study performed in the same Kenyan site in which no drug treatment was given during the vaccination phase (4–6,14). The most surprising result in our study was the lack of VE in the baseline *Pf*-negative cohort, although the individuals mounted the same level of anti-CS antibody titers post-vaccination as compared to those in the *Pf-*positive cohort, indicating that this is not due to vaccine non-responsiveness. Any comparison of differences between the baseline *Pf-*positive cohort with the baseline *Pf*-negative cohort must take into consideration that, 1) baseline *Pf* status represents a single point in time, 2) baseline *Pf* status is not randomly assigned and, 3) there was a small temporal delay in the recruitment of baseline *Pf*- positive compared to *Pf*-negative enrollees. One possible explanation is that baseline *Pf*- negative participants already have meaningful natural immunity against *Pf* infection and VE was masked by the stronger natural immunity. We are currently using trial samples to interrogate disparate acquired and innate immunity between these two cohorts at baseline. Another explanation is disparate exposure to *Pf*-infected mosquitoes; we were unable to show remarkable differences between these two cohorts upon geospatial analysis of domicile (Supplementary Figure S3). Given the caveats cited above, there may be other environmental/design factors contributing to the observed lack of VE in baseline *Pf*-negative cohort.

Antibody responses to CS protein and HBs were similar with or without drug treatment, with or without baseline *Pf* infection, and, most surprisingly, whether a *Pf* PCR+ event occurred during the ADI period. This is contrary to results from CHMI studies in malaria-naïve adults which showed that anti-CS antibodies are associated with protection (4,5,14,22). This suggests that, in the context of natural exposure with pre-existing immunity, the immunological ‘correlates of protection’ are more complicated than in CHMI studies in malaria naïve individuals.

This study has several limitations in identifying approaches that could be applied to overcome immune hyporesponsiveness and increase vaccine-induced protection against infection. First, the endpoint measurements were assessed only in groups that included the provision of antimalarial medications during vaccine doses. Second, we do not know whether adults are the most applicable targets for malaria elimination and whether this age group contributes heavily to ongoing transmission versus school-aged children and adolescents. Third, the design of the CS antigen component with the inclusion of multiple repeating units of the NANP epitope may adversely affect the induction of memory B cell responses critical for recall upon booster doses of vaccine. Next-generation CS-based vaccine candidates currently in preclinical development limit the number of NANP repeats or delete the C-term domain, hypothesized by some to be a decoy. These changes may increase the potency of a CS-based vaccine. Fourth, the dosage of RTS,S/AS01 used in this study was derived from earlier studies in malaria-naïve adults, and it may be necessary to use a higher dose in malaria-experienced adults, akin to the recommendation of high-dose influenza vaccines recommended for the elderly.

In conclusion, a vaccine comprising only the CS protein is unlikely to be sufficient to accelerate *Pf* malaria elimination due to modest efficacy in preventing infection, including in adults. This is likely due to the need for antibodies to neutralize the parasite in the short period of time prior to it entering the liver-stage development phase. Therefore, multi-antigen/multi-stage vaccine approaches that include a CS-component plus one or more blood-stage and/or sexual-stage (transmission-blocking) antigens, have risen to the top of future vaccine strategies.

## Data Availability

All data produced in the present work are contained in the manuscript

## Acknowledgements

We are grateful to the study volunteers for their participation in this clinical trial. We thank GlaxoSmithKline Biologicals SA for RTS,S/AS01_E_. We also acknowledge the following members of the study team, without whose hard work and dedication during a pandemic, this study would not have been possible: KEMRI (Walter Otieno, Eric Rono, Janet Oyieko, Valentine Sing’oei, Eliasa Bett, Dorothy Okello, Rachel Aguttu, Jacob Jagongo, Kennedy Otieno, Raymond Miyere, Roseline Apamo, Roseline Ohore, Alex Arika, Linah Ooro, Ludia Mbathi, Beatrice Orando, and the field team), PATH (Krystal Graham, Margaret Toher, Lionel Martellet, Linda Hoang, Trevor Lutzenhiser, Heather Richards, Richard Okwanyo, Shannon Shanahan, Rebecca Sanders, Kelsey Mertes, Allison Clifford, Kit Dubel, Lalaine Anova, Kerry Laurino), FHI-Clinical (Daniel Joffe, Michelle Pentikainen, Neo Choabi, Victorine Owira, Chantel Friend, Nicola van Zyl, Greg Tippett, Thando Madonsela, Monique Goldblatt, Loga Ganesh), DF/Net (Naydene Slabbert, Megan Baer, Gavin Robertson, Brian Postle, Joe Jiang) and GSK (Jasper Solomon, Pascale Vandoolaeghe, Jeroen Neirinck). We also thank Arianna Marini, Marija Zaric, Heejin Yun, Rachel Bromell, Morolayo Ayorinde, and Claire Streatfield at the Human Immunology Laboratory at the International AIDS Vaccine Initiative for the validation of the HBs antibody test and for generating the anti-HBs data for this study, and Kansas State Veterinary Diagnostic Laboratory for performance of the rabies antibody assay.

## Disclaimer

Material has been reviewed by the Walter Reed Army Institute of Research. There is no objection to its presentation and/or publication. The opinions or assertions contained herein are the private views of the author, and are not to be construed as official, or as reflecting true views of the Department of the Army or the Department of Defense. The investigators have adhered to the policies for protection of human subjects as prescribed in AR 70–25.

## Mention of any meeting(s) where the information has previously been presented

Posters presented at American Society for Tropical Medicine and Hygiene ASTMH 72nd Annual Meeting held at Hyatt Regency Chicago, Chicago IL, USA October 18-22, 2023 and the Multilateral Initiative on Malaria 9th Pan African Malaria Conference held in Kigali Convention Centre, Rwanda 21-27 April 2024.

## Change in author affiliations

JJA is currently at BioNTech, Mainz, Germany; LM is currently at Fred Hutchinson Cancer Research Center, Seattle, WA, USA; BA is currently at United States Center for Disease Control,

Nairobi, Kenya, LO, JDO, HA, AO are at Victoria Biomedical Research Institute, Kisumu, Kenya.

## Figure Legends

**Supplementary Table S1.** Participant randomization.

| Group | Vaccine | Sample size | Baseline<br>parasitemia | Anti-malarial<br>treatment | Rationale for<br>anti-malarial<br>treatment |
| --- | --- | --- | --- | --- | --- |
| 1 | RTS,S/AS01<sub>E</sub> | 164 | positive | yes | clear<br>parasites |
| 2 | RTS,S/AS01<sub>E</sub> | 128 | negative | yes | prophylaxis |
| 3 | RTS,S/AS01<sub>E</sub> | 35 | positive | no | Not treated |
| 4 | Rabies vaccine | 164 | positive | yes | clear<br>parasites |
| 5 | Rabies vaccine | 128 | negative | yes | prophylaxis |

**Supplementary Table S2.** Antimalarial drug treatment and vaccine schedule.

| Group #<sup>1</sup> | Month -1 | Month 0<br>(Vaccine 1) | 1-2 wks<br>before Vx<br>dose 2 | Month 1<br>(Vaccine 2) | 1 wk before<br>Vx dose 3 | Month 7<br>(Vaccine 3) | Month<br>8 to 14 |
| --- | --- | --- | --- | --- | --- | --- | --- |
| Group 1* | DHA/Pip +<br>LD PQ | RTS,S/AS01E | DHA/Pip +<br>LD PQ | RTS,S/AS01E | A/L + LD PQ | 1/5<sup>th</sup> dose<br>RTS,S/AS01E | ADI |
| Group 2* | DHA/Pip +<br>LD PQ | RTS,S/AS01E | DHA/Pip +<br>LD PQ | RTS,S/AS01E | A/L + LD PQ | 1/5<sup>th</sup> dose<br>RTS,S/AS01E | ADI |
| Group 3* | - | RTS,S/AS01E | - | RTS,S/AS01E | - | 1/5<sup>th</sup> dose<br>RTS,S/AS01E | - |
| Group 4 | DHA/Pip +<br>LD PQ | Rabies | DHA/Pip +<br>LD PQ | Rabies | A/L + LD PQ | Rabies | ADI |
| Group 5 | DHA/Pip +<br>LD PQ | Rabies | DHA/Pip +<br>LD PQ | Rabies | A/L + LD PQ | Rabies | ADI |

Vx: vaccine

**Supplementary Table S3.** Demographics.

| Parameter |  | Group 1 | Group 2 | Group 3 | Group 4 | Group 5 |
| --- | --- | --- | --- | --- | --- | --- |
| Total Vaccinated Cohort, n |  | 160 | 124 | 35 | 159 | 127 |
| Age, mean (SD) |  | 31.94 (8.04) | 34.02 (7.63) | 34.57 (8.93) | 31.76 (8.49) | 34.04 (8.04) |
| Sex (n [%]) | Female | 72 (45.0) | 80 (64.5) | 21 (60.0) | 75 (47.2) | 75 (59.1) |
|  | Male | 88 (55.0) | 44 (35.5) | 14 (40.0) | 84 (52.8) | 52 (40.9) |
| Ethnicity | Luo | 158 (98.8) | 122 (98.4) | 35 (100) | 159 (100) | 125 (98.4) |
|  | Luhya | 2 (1.3) | 2 (1.6) | 0 | 0 | 2 (1.6) |
| HIV Status (n [%]) | Positive | 23 (14.4) | 24 (19.4) | 7 (20.0) | 24 (15.1) | 25 (19.7) |
|  | Negative | 137 (85.6) | 100 (80.6) | 28 (80.0) | 135 (84.9) | 102 (80.3) |
| Bednet ownership (n [%]) | Yes | 134 (83.8) | 106 (85.5) | 33 (94.3) | 133 (83.6) | 101 (79.5) |
|  | No | 26 (16.3) | 18 (14.5) | 2 (5.7) | 26 (16.4) | 26 (20.5) |
| Sleep Under Net n [%]) | Yes | 134 (83.8) | 106 (85.5) | 33 (94.3) | 133 (83.6) | 101 (79.5) |
|  | Daily | 134 (83.8) | 106 (85.5) | 33 (94.3) | 133 (83.6) | 101 (79.5) |
| IRS* n [%]) | Yes | 3 (1.9) | 2 (1.6) | 1 (2.9) | 3 (1.9) | 4 (3.1) |
|  | No | 157 (98.1) | 122 (98.4) | 34 (97.1) | 156 (98.1) | 123 (96.9) |
| * IRS, Indoor residual spray within the past year n [%]) |  |  |  |  |  |  |

n: number of participants
SD: standard deviation
IRS: indoor residual spray within the past year

**Supplementary Table S4.** Safety.

|  |  | RTS,S/AS01E |  |  | Rabies |  | Total<br>(N=605)<br>n (%) |
| --- | --- | --- | --- | --- | --- | --- | --- |
|  |  | Group 1<br>(N=160)<br>n (%) | Group 2<br>(N=124)<br>n (%) | Group 3<br>(N=35)<br>n (%) | Group 4<br>(N=159)<br>n (%) | Group 5<br>(N=127)<br>n (%) |  |
| **Local Solicited AEs** | Number of AEs | 50 | 58 | 34 | 0 | 0 | 142 |
|  | Number of Participants | 24 (49.0) | 26 (55.3) | 14 (40.0) | 0 | 0 | 64 (48.9) |
| **Systemic Solicited AEs** | Number of AEs | 44 | 47 | 33 | 0 | 0 | 124 |
|  | Number of Participants | 20 (40.8) | 16 (34.0) | 14 (40.0) | 0 | 0 | 50 (38.2) |
| **Unsolicited AEs** | Number of AEs | 184 | 178 | 28 | 223 | 186 | 799 |
|  | Number of Participants | 103 (64.4) | 87 (70.2) | 20 (57.1) | 110 (69.2) | 92 (72.4) | 412 (68.1) |
| **SAEs** | Number of AEs | 3 | 4 | 1 | 1 | 4 | 13 |
|  | Number of Participants | 3 (1.9) | 2 (1.6) | 1 (2.9) | 1 (0.6) | 3 (2.4) | 10 (1.7) |

Rabies: rabies vaccine
N: Total number of participants vaccinated
n: Number of participants with event
AEs: adverse events
SAE: serious adverse events

**Supplementary Table S5.** Anti-NANP avidity indices.

| Visit number | Statistics | Group 1 | Group 2 | Group 3 |
| --- | --- | --- | --- | --- |
| Visit 2 - Day -27 | Mean | 0.59 | 0.64 | 0.66 |
|  | LL, UL | 0.55, 0.63 | 0.60, 0.68 | 0.58, 0.73 |
|  | Min, Max | 0.12, 0.84 | 0.09, 0.90 | 0.13, 0.88 |
|  | p-value | 1 vs 2 (0.182) | 2 vs 3 (0.708) | 3 vs 1 (0.196) |
| Baseline | Mean | 0.62 | 0.64 | 0.65 |
|  | LL, UL | 0.58, 0.65 | 0.60, 0.68 | 0.57, 0.73 |
|  | Min, Max | 0.10, 0.96 | 0.10, 0.84 | 0.16, 0.89 |
|  | p-value | 1 vs 2 (0.417) | 2 vs 3 (0.458) | 3 vs 1 (0.185) |
| Visit 11 - Day 29 | Mean | 0.64 | 0.67 | 0.69 |
|  | LL, UL | 0.62, 0.67 | 0.65, 0.70 | 0.62, 0.76 |
|  | Min, Max | 0.16, 1.01 | 0.28, 1.00 | 0.32, 1.35 |
|  | p-value | 1 vs 2 (0.180) | 2 vs 3 (0.521) | 3 vs 1 (0.129) |
| Visit 13 - Day 57 | Mean | 0.63 | 0.64 | 0.67 |
|  | LL, UL | 0.60, 0.65 | 0.61, 0.67 | 0.62, 0.72 |
|  | Min, Max | 0.26, 1.23 | 0.19, 1.43 | 0.44, 1.08 |
|  | p-value | 1 vs 2 (0.496) | 2 vs 3 (0.289) | 3 vs 1 (0.128) |
| Visit 17 - Day 197 | Mean | 0.59 | 0.63 | 0.66 |
|  | LL, UL | 0.57, 0.62 | 0.60, 0.66 | 0.60, 0.71 |
|  | Min, Max | 0.10, 1.05 | 0.17, 0.94 | 0.35, 0.98 |
|  | p-value | 1 vs 2 (0.061) | 2 vs 3 (0.368) | 3 vs 1 (0.033) |
| Visit 20 - Day 225 | Mean | 0.55 | 0.65 | 0.66 |
|  | LL, UL | 0.53, 0.58 | 0.62, 0.68 | 0.60, 0.72 |
|  | Min, Max | 0.11, 0.94 | 0.19, 1.26 | 0.27, 1.20 |
|  | p-value | 1 vs 2 (0.000) | 2 vs 3 (0.747) | 3 vs 1 (0.000) |
| Visit 27 - Day 372 | Mean | 0.52 | 0.57 | 0.55 |
|  | LL, UL | 0.49, 0.55 | 0.54, 0.60 | 0.49, 0.62 |
|  | Min, Max | 0.09, 0.86 | 0.17, 0.90 | 0.26, 0.89 |

|  | p-value | 1 vs 2 (0.011) | 2 vs 3 (0.846) | 3 vs 1 (0.147) |
| --- | --- | --- | --- | --- |
| Visit 39 (End of Study) | Mean | 0.53 | 0.55 | 0.56 |
|  | LL, UL | 0.50, 0.55 | 0.52, 0.58 | 0.49, 0.62 |
|  | Min, Max | 0.14, 0.85 | 0.23, 0.87 | 0.27, 0.88 |
|  | p-value | 1 vs 2 (0.325) | 2 vs 3 (0.873) | 3 vs 1 (0.425) |

LL: lower limit
UL: upper limit
Min: minimum
Max: maximum

**Supplementary Table S6.**
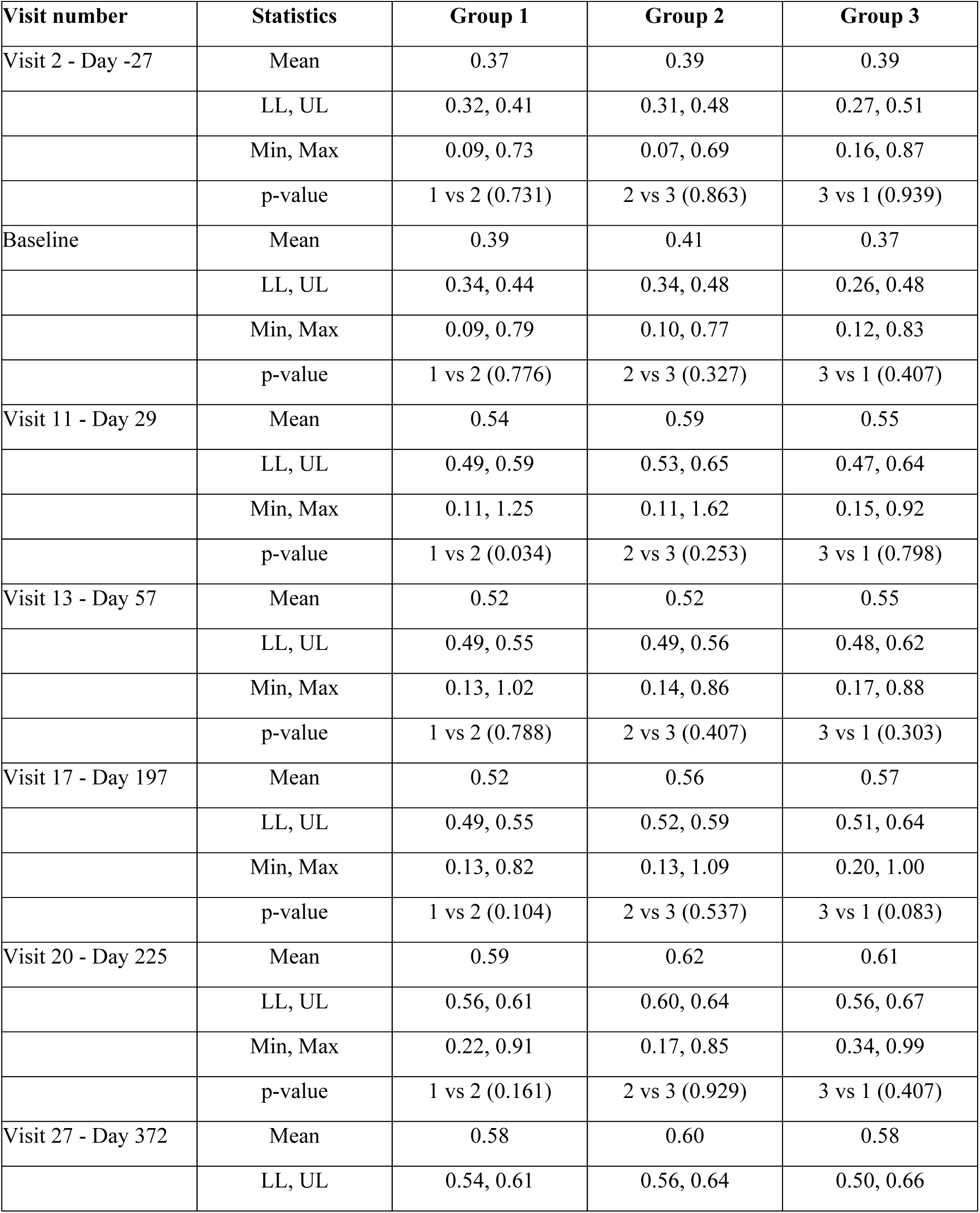
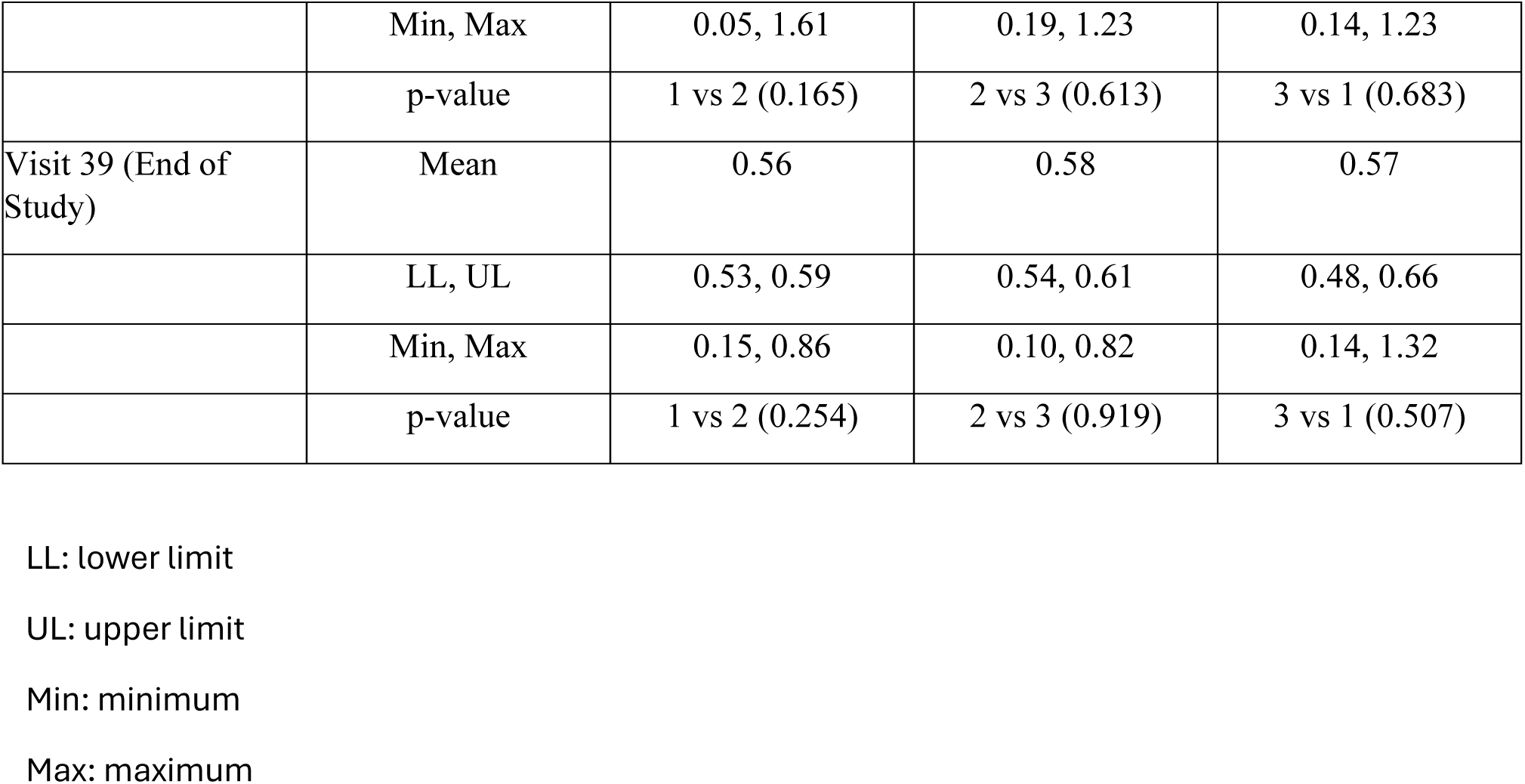
Anti-C-term avidity indices.

| Visit number | Statistics | Group 1 | Group 2 | Group 3 |
| --- | --- | --- | --- | --- |
| Visit 2 - Day -27 | Mean | 0.37 | 0.39 | 0.39 |
|  | LL, UL | 0.32, 0.41 | 0.31, 0.48 | 0.27, 0.51 |
|  | Min, Max | 0.09, 0.73 | 0.07, 0.69 | 0.16, 0.87 |
|  | p-value | 1 vs 2 (0.731) | 2 vs 3 (0.863) | 3 vs 1 (0.939) |
| Baseline | Mean | 0.39 | 0.41 | 0.37 |
|  | LL, UL | 0.34, 0.44 | 0.34, 0.48 | 0.26, 0.48 |
|  | Min, Max | 0.09, 0.79 | 0.10, 0.77 | 0.12, 0.83 |
|  | p-value | 1 vs 2 (0.776) | 2 vs 3 (0.327) | 3 vs 1 (0.407) |
| Visit 11 - Day 29 | Mean | 0.54 | 0.59 | 0.55 |
|  | LL, UL | 0.49, 0.59 | 0.53, 0.65 | 0.47, 0.64 |
|  | Min, Max | 0.11, 1.25 | 0.11, 1.62 | 0.15, 0.92 |
|  | p-value | 1 vs 2 (0.034) | 2 vs 3 (0.253) | 3 vs 1 (0.798) |
| Visit 13 - Day 57 | Mean | 0.52 | 0.52 | 0.55 |
|  | LL, UL | 0.49, 0.55 | 0.49, 0.56 | 0.48, 0.62 |
|  | Min, Max | 0.13, 1.02 | 0.14, 0.86 | 0.17, 0.88 |
|  | p-value | 1 vs 2 (0.788) | 2 vs 3 (0.407) | 3 vs 1 (0.303) |
| Visit 17 - Day 197 | Mean | 0.52 | 0.56 | 0.57 |
|  | LL, UL | 0.49, 0.55 | 0.52, 0.59 | 0.51, 0.64 |
|  | Min, Max | 0.13, 0.82 | 0.13, 1.09 | 0.20, 1.00 |
|  | p-value | 1 vs 2 (0.104) | 2 vs 3 (0.537) | 3 vs 1 (0.083) |
| Visit 20 - Day 225 | Mean | 0.59 | 0.62 | 0.61 |
|  | LL, UL | 0.56, 0.61 | 0.60, 0.64 | 0.56, 0.67 |
|  | Min, Max | 0.22, 0.91 | 0.17, 0.85 | 0.34, 0.99 |
|  | p-value | 1 vs 2 (0.161) | 2 vs 3 (0.929) | 3 vs 1 (0.407) |
| Visit 27 - Day 372 | Mean | 0.58 | 0.60 | 0.58 |
|  | LL, UL | 0.54, 0.61 | 0.56, 0.64 | 0.50, 0.66 |
|  | Min, Max | 0.05, 1.61 | 0.19, 1.23 | 0.14, 1.23 |
|  | p-value | 1 vs 2 (0.165) | 2 vs 3 (0.613) | 3 vs 1 (0.683) |
| Visit 39 (End of Study) | Mean | 0.56 | 0.58 | 0.57 |
|  | LL, UL | 0.53, 0.59 | 0.54, 0.61 | 0.48, 0.66 |
|  | Min, Max | 0.15, 0.86 | 0.10, 0.82 | 0.14, 1.32 |
|  | p-value | 1 vs 2 (0.254) | 2 vs 3 (0.919) | 3 vs 1 (0.507) |

LL: lower limit
UL: upper limit
Min: minimum
Max: maximum

**Figure S1.**
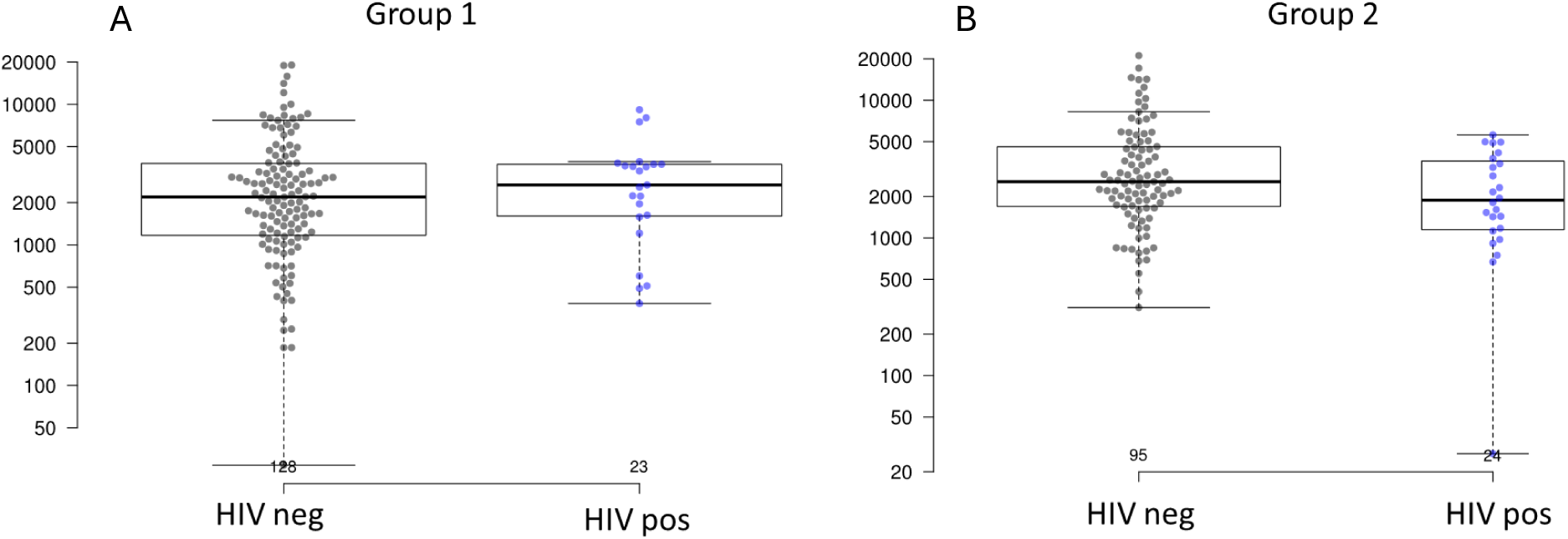
Anti-NANP titers in HIV-negative and HIV-positive participants in Groups 1 (A) and 2 (B). Abbreviations: EU, ELISA units, equivalent to Geometric Mean Titers, Pos, positive; Neg, negative.

**Figure S2.**
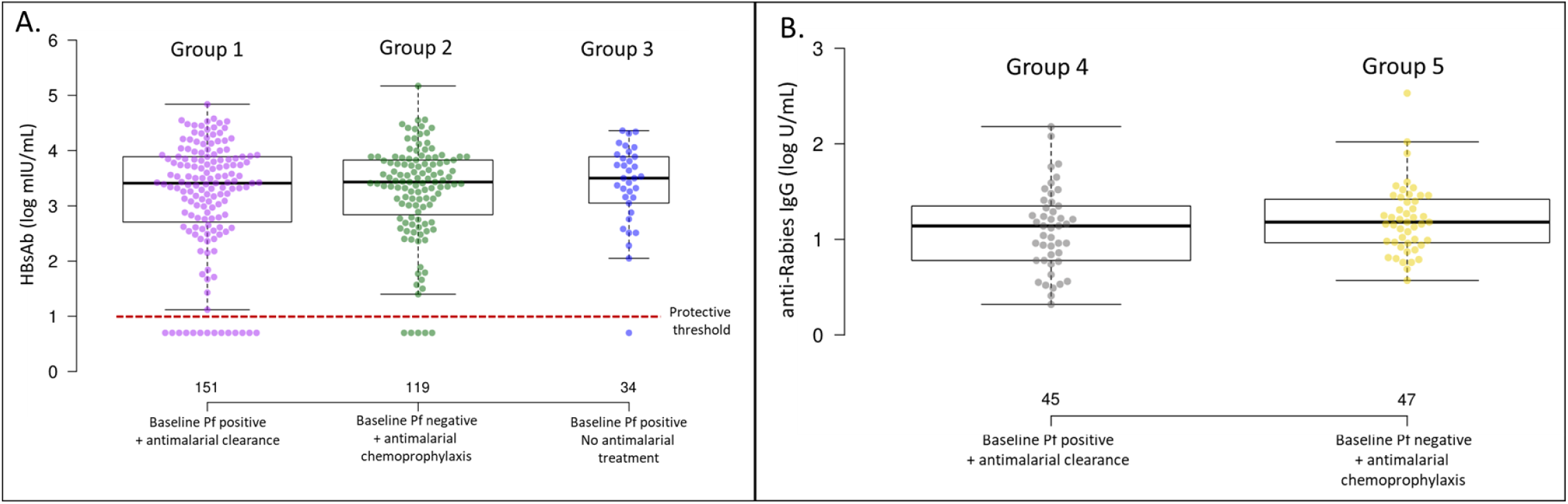
Anti-hepatitis B surface antigen and anti-rabies antibody concentrations post-dose 3 (day 225). A. Anti-HBs titers in Groups 1, 2 and 3. B. Anti-rabies titers in Groups 4 and 5. Abbreviations: HBs, hepatitis B surface antigen; mIU/mL, milli-international units per milliliter; *Pf*, *Plasmodium falciparum*; U/mL, units per milliliter.

**Figure S3.**
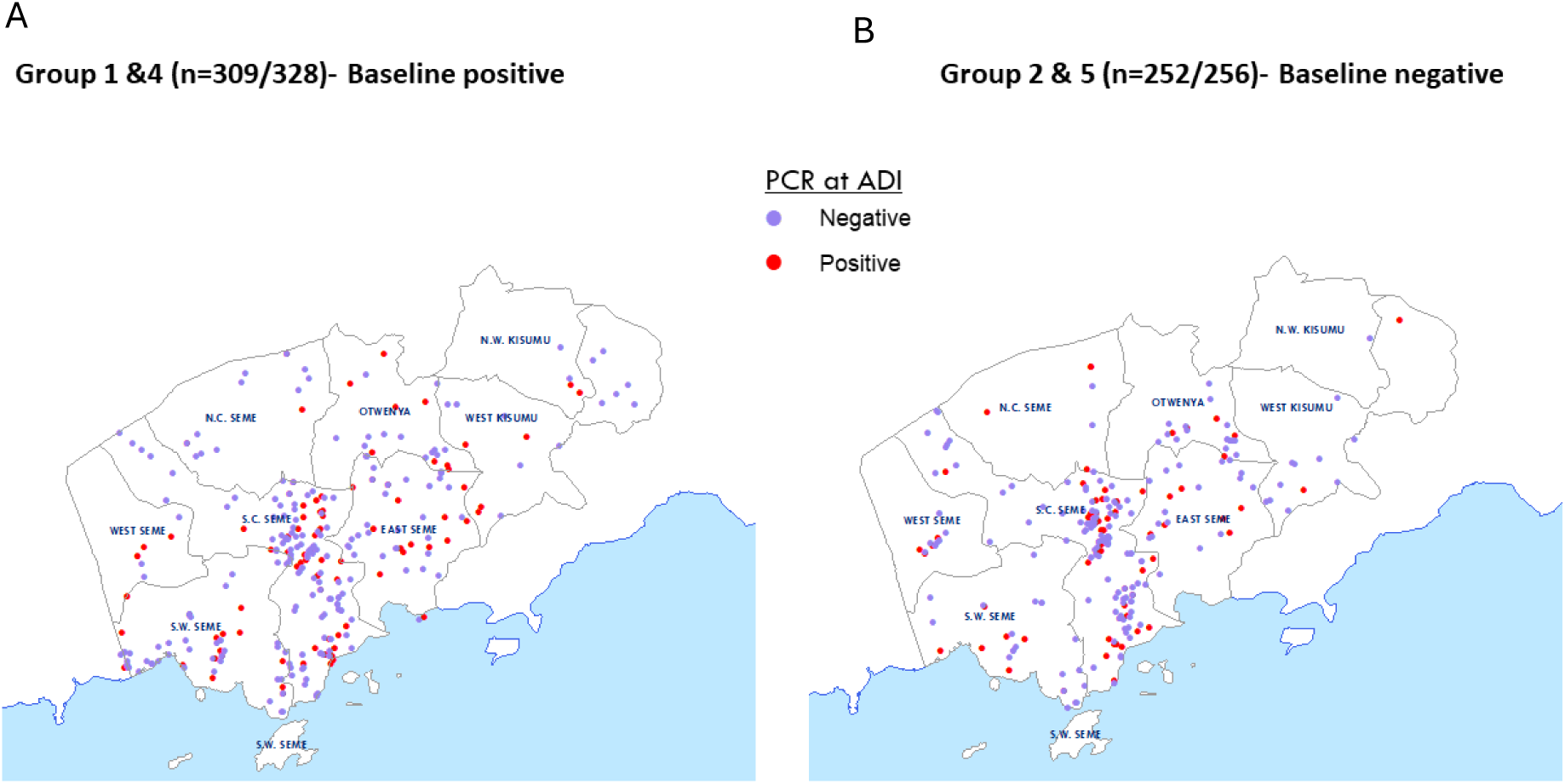
Geospatial mapping of participants within the study area. A. Domicile of baseline Pf-positive participants in Groups 1 and 4 compared to, B. domicile of baseline Pf-negative participants in Groups 2 and 5 broken out by whether they had a *Pf* PCR positive event during active detection of infection phase. Abbreviations: ADI, active detection of infection; *Pf*, *Plasmodium falciparum*.

